# Comparison of SARS-CoV-2 serological tests with different antigen targets

**DOI:** 10.1101/2020.07.09.20149864

**Authors:** Alix T. Coste, Katia Jaton, Matthaios Papadimitriou-Olivgeris, Gilbert Greub, Antony Croxatto

**Affiliations:** Institute of Microbiology, Lausanne University Hospital and University of Lausanne, Lausanne, Switzerland; Service of Hospital Preventive Medicine, Lausanne University Hospital, Switzerland; Service of Infectious Diseases, Lausanne University Hospital, Switzerland

**Keywords:** SARS-CoV-2, serology, evaluation, Kits

## Abstract

**Background:** These last months, dozens of SARS-CoV-2 serological tests have become available with varying performances. A major effort was completed to compare 17 serological tests.

**Methods:** In a preliminary phase, we compared 17 IgG, IgM, IgA and pan Ig serological tests including ELISA, LFA, CLIA and ECLIA on a panel of 182 sera, comprising 113 sera from hospitalized patients with a positive RT-PCR, and 69 sampled before 1^st^ November 2019, expected to give a positive and negative results, respectively. In a second phase, the five best performing and most available tests were further evaluated on a total of 582 sera (178 and 404 expected positive and negative, respectively), allowing the assessment of 20 possible cross-reactions with other virus.

**Results:** In the preliminary phase, among eight IgG/pan-Ig ELISA or CLIA/ECLIA tests, four had a sensitivity and specificity above 90% and 98% respectively, and on six IgM/IgA tests, only one was acceptable. Only one LFA test on three showed good performances for both IgG and IgM. For all the tests IgM and IgG aroused concomitantly. In the second phase, no tests showed particular cross-reaction. We observed an important heterogeneity in the development of the antibody response, and that anti-nucleocapside (anti-N) antibodies appeared earlier than the anti-spike (anti-S) proteins.

**Conclusions:** The identified SARS-CoV-2 serology tests may be used for the diagnostic of CoviD-19 for negative RT-PCR patients presenting severe to mild suggestive symptoms or particular clinical presentation. Detection of both anti-N and anti-S could be complementary to increase the sensitivity of the analysis.

## Introduction

In December 2019, a new virus causing severe respiratory infections emerged in China in the Wuhan area. This virus was named SARS-CoV-2 (Severe Acute Respiratory Syndrome Coronavirus 2) and the associated disease was coined “COVID-19” (COronaVIrus Disease 2019). The epidemic rapidly spread and the WHO classified it as a pandemic in March 2020 (https://www.who.int/news-room/detail/27-04-2020-who-timeline---covid-19). This new virus was classified in the Coronaviridae family and in the *Betacoronavirus* genus like the SARS-CoV and the MERS-CoV (Middle East Respiratory Syndrome Coronavirus).

The mortality rate of the SARS-CoV-2 (about 2%) is lower than SARS-CoV and MERS-CoV (10 and 30%, respectively) but its reproduction rate R0 (2 to 2.5) is higher, than the SARS-CoV (1.7 to 1.9) and the MERS-CoV (<1), probably explaining its rapid spreading worldwide [1-3]. In a first phase of the pandemic, nucleic acid amplification tests (NAAT) enabled rapid detection of infected patients, their sorting and their possible isolation. In a second phase, serology testing appeared particularly important as it permits to diagnose patients after the acute phase of the infection or with atypical clinical presentation with no nasopharyngeal shedding of the virus [4, 5]. Indeed, in contrast to NAAT, which must be carried out when and where the virus is excreted, the serological assays might be performed anytime ideally more than two weeks after symptoms onset. Serology also appeared to be the tests of choice to perform large-scale population prevalence studies.

Various SARS-CoV-2 serological tests using different antigenic proteins have been arriving on the market the last two-three months (https://www.finddx.org/covid-19/pipeline). Some of them use whole virus lysate, recombinant full S (spike) or N (nucleocapsid) proteins, peptides of the N or specific domains S1, S2 or RBD (receptor-binding domains) of the S protein.

Different studies demonstrated that the S and N proteins were the most immunogenic [6]. The N protein is relatively small with no glycosylated sites and presents a higher level of conservation than the S protein among coronavirus infecting human, allowing possible false positive results through cross-reaction [4, 7, 8]. In contrast the S protein is a large transmembrane protein, less conserved, containing several glycosylated sites and bearing a more complex conformation, leading to production of more specific antibodies often recognizing conformational or glycosylated epitopes [7-9]. Thus the use of recombinant S protein lacking glycosylation or conformation in immunoassays may lead to false negative results.

In this study we evaluated several SARS-CoV-2 serological tests including, ELISA (Enzyme-Linked ImmunoSorbent Assays), LFA (Lateral Flow ImmunoAssays), CLIA (ChemiLuminescent ImmunoAssays) or ECLIA (Electro-ChemiLuminescent ImmunoAssay). This evaluation is primarily aiming to identify high quality tests for symptomatic patients but also to guide other diagnostic laboratories.

## Material and Methods

### Samples

The first phase of the evaluation was performed on 182 sera (113 positive and 69 negative) (Supplementary Table S1). Then, the evaluation was completed for the selected tests on 400 sera (65 positive and 335 negative), leading to a full evaluation performed on 582 sera (178 positive and 404 negative) (Supplementary Table S1). Negative-expected sera were selected among sera sampled before the 1^st^ November 2019 and indicated as “Anterior” for anterior to SARS-CoV-2 pandemic (Table 1). Possible cross-reactivity was assesses through testing of sera known to be positive for a given microorganism, indicated as “Anterior (microorganism)”. The positive-expected sera were sampled during the first 2 months post-symptoms from patients documented with a positive SARS-CoV-2 RT-PCR. The dates of symptoms were extracted from the electronic records of the SARS-CoV-2 RT-PCR patients for the 178 positive sera.

### ELISA, LFA, and CLIA assays

Each test (Supplementary Table S2) was performed according to the manufacturers’ instructions. ELISA assays were done in duplicates and manually to diminish dead volume, except washing steps performed with a microplate washer (PW40, Bio-Rad, France). Lecture of the Optical densities (OD) was done with a microplate reader (800 TSI, BioTek, USA). For CLIA assays, the LIAISON^®^ SARS-CoV-2 IgG kit was performed on a Liaison^®^ XL (Diasorin, Italy), and the MAGLUMI™ 2019-nCoV IgG and IgM kits on a MAGLUMI™ 800 (Snibe, China). The ECLIA assay, Elecsys anti-SARS-CoV-2 was performed on a COBAS 6000 (Roche, Switzerland).

Sensitivity was evaluated on expected positive sera according to day post-symptoms. Specificity was determined on expected negative sera sampled before November 1^st^ 2019.

### Statistical analyses

Sensitivity and specificity with 95% CI (Wilson/Brown method of GraphPad Prism 8.3.0) were calculated with Excel and GraphPad prims.

## Results

### Preliminary evaluation of 17 SARS-CoV-2 serologic tests

A preliminary evaluation of 17 serological kits (Table S2) has been performed on 182 sera, including 113 sera from patients positive for a SARS-CoV-2 RT-PCR (considered as positive) and 69 sera sampled before November 1, 2019, (considered as negative). For the 113 so-called “positive sera”, a stratification of the results was done according to the time between symptoms onset and sera sampling. Four categories were defined 0-5 days, 6-10 days, 11-15 days and >15 days. The 17 serological kits tested included 10 ELISA (five IgG, three IgM, one IgA, and one IgM+IgA) from five manufacturers, three LFA (IgG+IgM) from three manufacturers, three CLIA (two IgG and one IgM) from two manufacturers, and one ECLIA (pan-Ig).

For all the 17 tests (IgG, IgM, IgA, pan-Ig) the sensitivity increased over time post-symptoms as expected (Figure 1, Table S3-5). Concerning IgG or pan-Ig tests, a sensitivity above 70 % was obtained after 10 days post-symptoms for almost all tests except the Diasorin ISON^®^ SARS-CoV-2 IgG kit (57%). However, a sampling at minimum 15 days post-symptoms is necessary for most of the IgG/pan-Ig tests to reach more than 90% sensitivity (Figure 1 and 2, Table S3-5). Only three tests exhibited a sensitivity lower than 90% more than 15 days post-symptoms, the Euroimmun ELISA IgG test (88%; CI: 72-95), the NADAL^®^ COVID-19 IgG/IgM LFA test (84 %; 95% CI: 67-93) and the Diasorin ISON^®^ SARS-CoV-2 IgG CLIA kit (83%; 95%CI: 66-93) (Figure 2; Table S3-5). All the tests except the SARS-CoV-2 NP IgG ELISA Kit from ImmunoDiagnostics limited presented a specificity equal or above 97%. Noteworthy, none of the IgG tests have shown specific cross reactivity with sera from patients documented with a positive RT-PCR for Human seasonal coronavirus (Hcov) E229, OC43, HKU1, and NL63.

**Figure 1:**
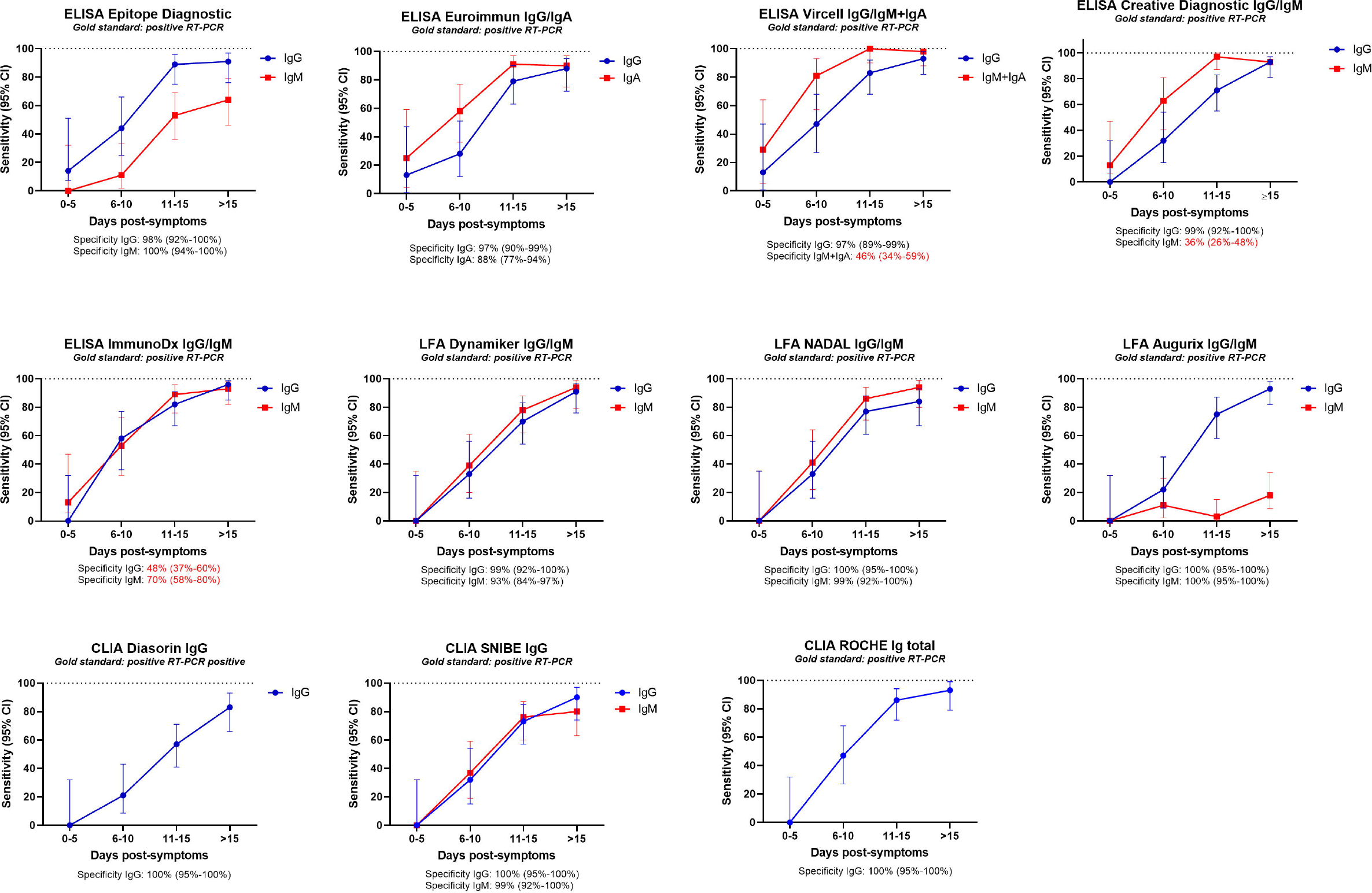
Preliminary evaluation: Sensitivity at 0-5, 6-10, 11-15 and above 15 days post-symptoms. Specificity is indicated below each graph. Poor specificities are in red characters.

**Figure 2:**
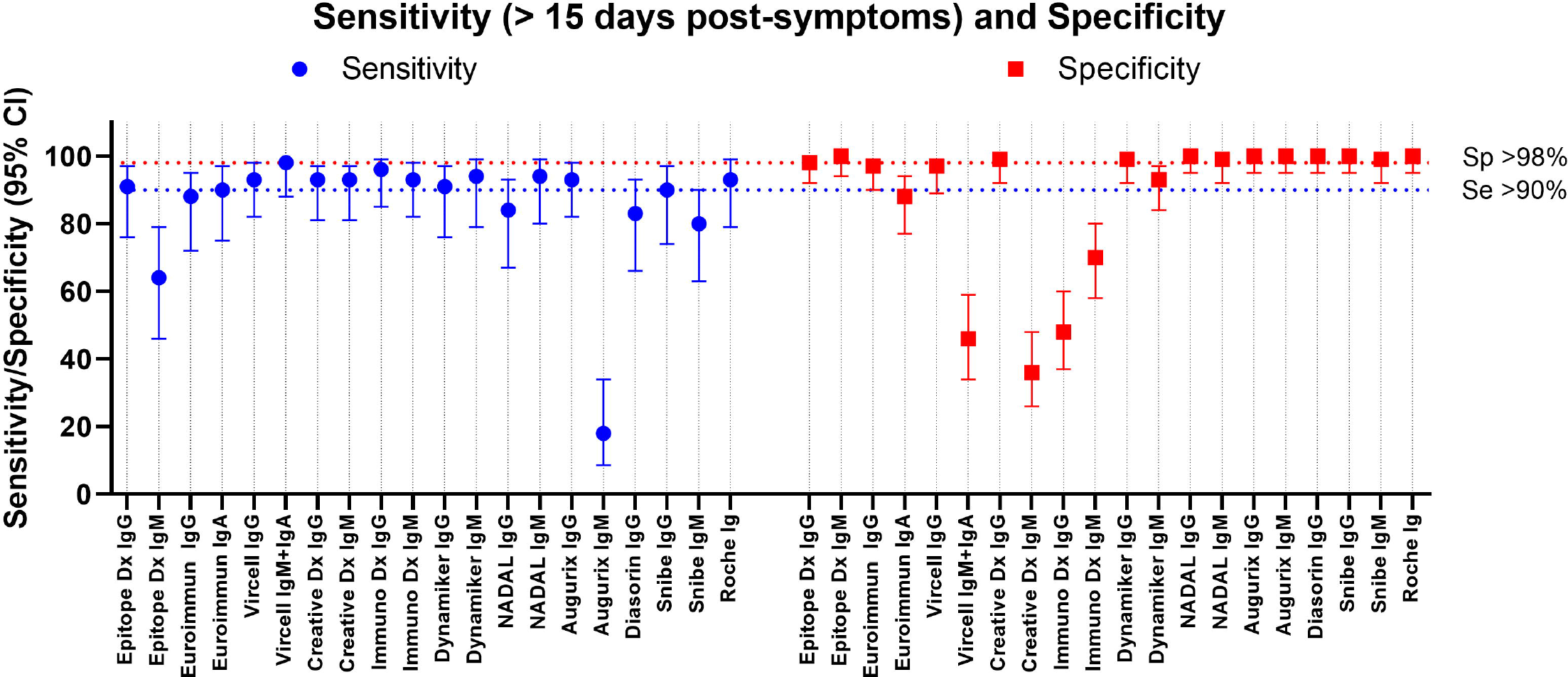
Preliminary evaluation. Comparison of the sensitivity and specificity. The sensitivity is given for the sample above 15 days post symptoms.

The only IgM/IgA tests performed at least 15 days from symptoms onset, exhibiting satisfying performances were the NADAL LFA (sensitivity 95%, 95% CI:80-99; specificity: 99%, 95% CI: 92-100), the Dynamiker LFA (sensitivity 94%, 95% CI:79-99; specificity: 93%, 95% CI: 84-97), and the CLIA from Snibe (sensitivity 80%, 95% CI:63-90; specificity: 99%, 95% CI: 92-100) (Figure 2; Table S3-5).

The other tests, Epitope Diagnostic (ED) IgM ELISA, Euroimmun IgA ELISA, Vircell IgM+IgA ELISA, ImmunoDiagnostic limited IgM ELISA demonstrated insufficient performances with sensitivity and specificity below 80% and 98%, respectively (Figure 2, Table S3-5).

Concerning the LFA, IgG and IgM being tested simultaneously, both tests should give excellent results to be valuable. The Dynamiker IgG/IgM LFA is the only test respecting a sensitivity and specificity of more than 90% for both Ig.

Interestingly, for the tests with an IgM specificity above 90% (Dynamiker LFA, NADAL LFA and Snibe CLIA), we observed a simultaneous IgM and IgG response overtime (Figure 1).

### Complete evaluation of 5 SARS-CoV-2 selected serologic tests

Following the preliminary evaluation, the ED IgG ELISA test, the Dynamiker IgG/IgM LFA test, the Diasorin IgG CLIA and the Snibe IgG and IgM CLIA tests were thus selected for further analyses based on i) sensitivity and specificity performance of the preliminary evaluation, ii) Protein detected (anti-N: ED IgG ELISA and Dynamiker IgG/IgM; anti-S: Diasorin IgG CLIA, anti N+S: Snibe IgG/IgM CLIA) iii) availability of the kits at the later on 15^th^ April 2020 in Switzerland, iv) specific detection of IgG and/or IgM or IgA and v) compatibility of the kits to most laboratory needs including median to low samples volumes per day and extended expiration days upon kits opening. For instance, despite its good performance, the ECLIA from Roche was not selected as it detects pan-Ig, which is not the most appropriate for infectious serology diagnostic.

All the five selected tests were further evaluated on 65 positive and 335 negative sera to end-up with a global evaluation performed on 178 positive and 404 negative for a total of 586 sera (Figure 3, Table S1). The negative sera were chosen to assess possible cross-reactivity with human viral infection other than human coronavirus: Herpes simplex virus 1 and 2, Respiratory Syncytial Virus, Epstein-Barr virus, Cytomegalovirus, Mumps virus, Measles virus, Parvovirus B19, Rubella virus, Tick-borne encephalitis virus, Influenza A and B, Varicella-zoster virus, Human Immunodeficiency virus, Hepatitis virus A, B, C, D, and E, and some rheumatoid factors, or auto-antibodies (anti-PR3, -PR4, SCL70, SCL71).

**Figure 3:**
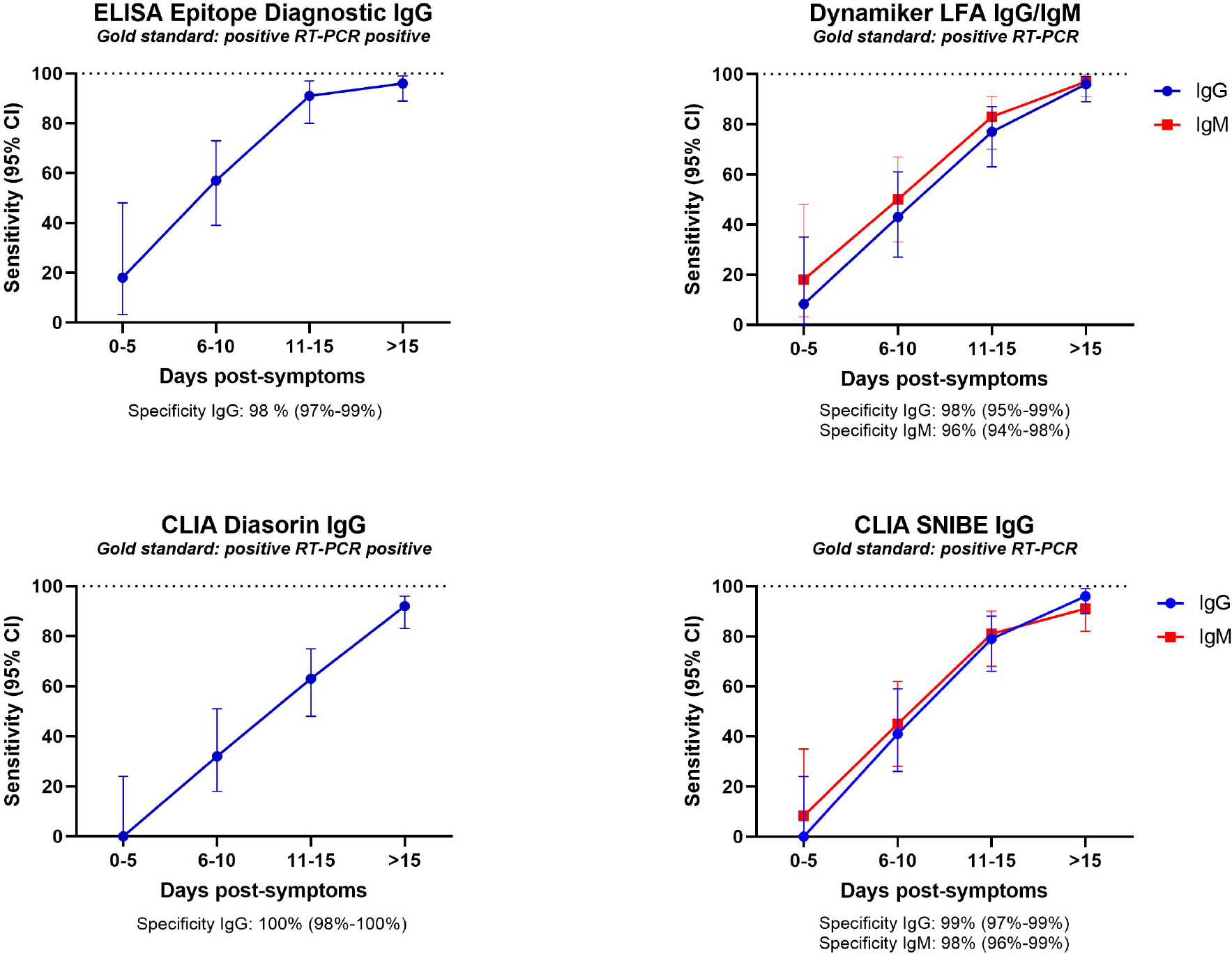
Complete evaluation: Sensitivity at 0-5, 6-10, 11-15 and above 15 days post-symptoms. Specificity is indicated below each graph.

The four IgG tests demonstrated good sensitivity (≥96%) and specificity (≥98%) performances at more than 15 days post-symptoms, except the Diasorin ISON^®^ SARS-CoV-2 IgG CLIA kit that showed a sensitivity of 92% (95% CI 83-96) but with a specificity of 100% (Figure 4 and Table S6). The IgM tests exhibited a sensitivity of 91% (95% CI 95-92), and a specificity of 98% (95% CI 96-99) for the CLIA Snibe IgM test with, and a sensitivity of 97 % (95% CI 91-100) and a specificity of 96% (95% CI 94-98) for the LFA Dynamiker IgM test (Figure 4 and Table S6).

**Figure 4:**
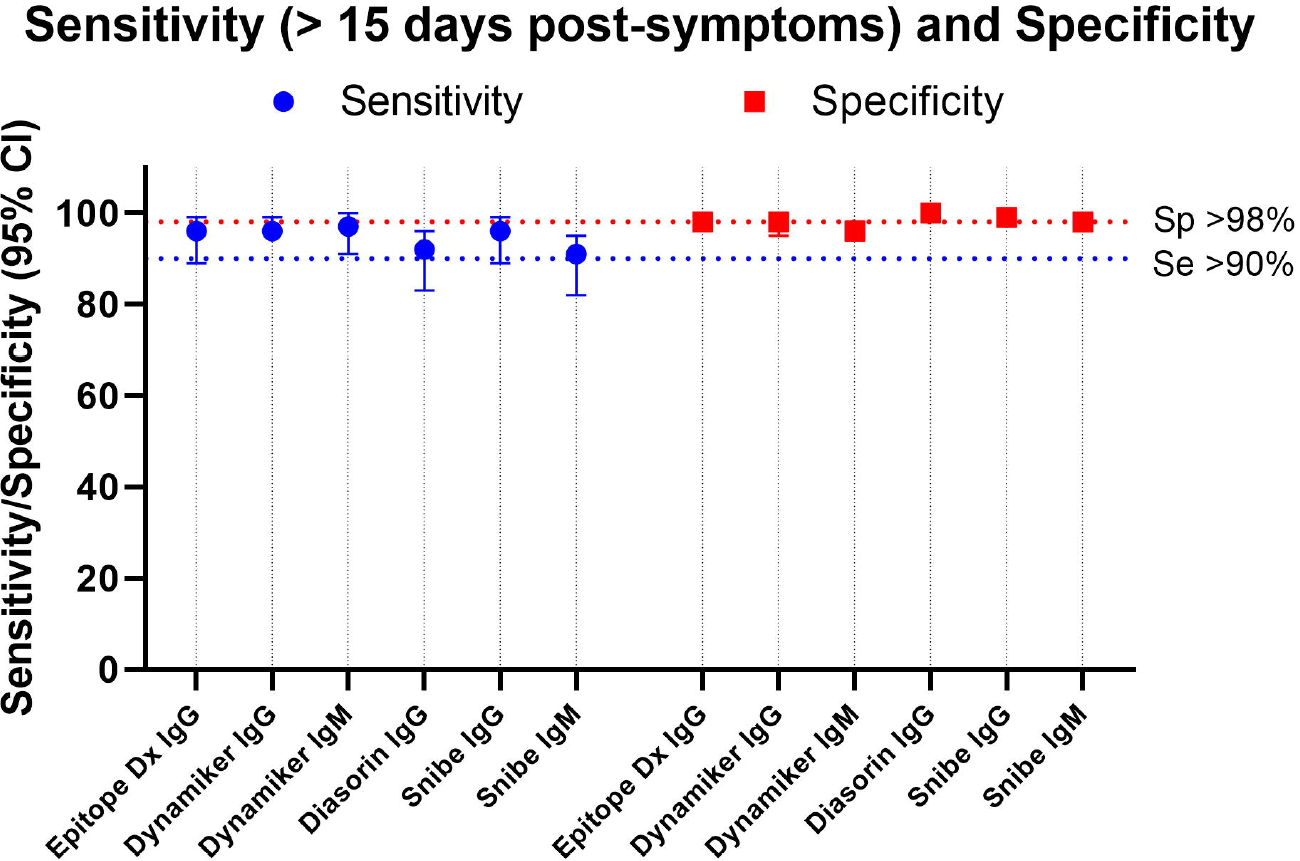
Complete evaluation: Comparison of the sensitivity and specificity. The sensitivity is given for the sample above 15 days post symptoms.

### Semi-quantitative antibody production

We analyzed the development of the IgG semi-quantitative response overtime post-symptoms with the ELISA from ED targeting the anti-N response, the CLIA from Diasorin targeting the anti-S1-S2 domains of the protein S, and the CLIA from Snibe targeting the anti-N and anti-S response (Figure 5). Among the 184 positive sera, several were sampled from the same patient overtime. We could observe that one patient with ELISA ED and two patients with CLIA from Diasorin and from Snibe became positive for anti-SARS-CoV-2 IgG more than 15 days post-symptoms (Figure 5). Thus, one patient was negative with the CLIA from Diasorin for two consecutive sera collected at days 17 and 21 post-symptoms and became positive only at day 27 post-symptoms. Similarly, one patient was negative with the CLIA from Snibe with the sera collected at days 13 post-symptoms and became positive only at day 21 post-symptoms. One patient became positive with all test only 32 days post-symptoms. Interestingly, we observed that the IgG anti-N response (ELISA ED) appeared more rapidly (Figure 5A and D) than the IgG anti-S response (CLIA Diasorin) (Figure 5B and D). Rationally, the response observed with the CLIA from SNIBE targeting both N and S protein appeared to be intermediate (Figure 5C).

**Figure 5:**
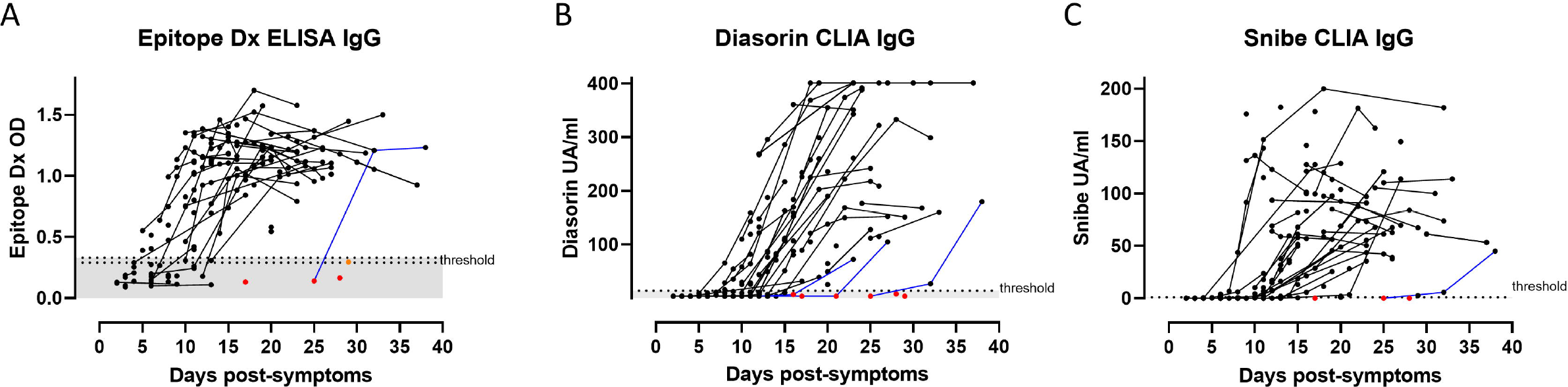
Kinetic of antibody production per patients. False negative results above 15 days post-symptoms are indicated with red dots and borderline results with orange dots. Patients with several collected sera are represented by connecting lines. Blue lines represent patients that became positive for anti-SARS-CoV-2 IgG more than 15 days post-symptoms.

## Discussion

To our knowledge, this study is the first one comparing so many different serologic tests for SARS-CoV-2 diagnostic using different technologies, LFA, ELISA, CLIA and ECLIA, on sera from RT-PCR positive patients collected over one to two months post-symptoms, and assessing 20 possible cross reaction with other viral infections.

This large evaluation of 17 SARS-CoV-2 serological tests highlights in the preliminary phase that among eight IgG/pan-Ig ELISA or CLIA/ECLIA tests, four were recommended with a combined sensitivity above 90% and specificity above 98%, and that on six IgM/IgA ELISA or CLIA/ECLIA tests, only one was acceptable with a combined sensitivity above 80% and specificity of 99%.

Concerning LFA, only one test showed good performances for both IgG and IgM, showing that a thorough evaluation is absolutely required before their commercialization in pharmacy or their use by private practitioners outside referenced diagnostic laboratories. For all different tests, we could observed that IgM and IgG aroused concomitantly, as already described for SARS infection [10]. This concomitant production of IgM and IgG together with the poor IgM performance of most tests suggests that the use of IgM is useless for the sero-diagnostic of SARS-CoV-2 acute/subacute infection. SARS-CoV-2 RT-PCR remains the test of choice for early diagnostic a few days after symptoms onset [11]. Whether IgM may be linked to the severity of the disease and may be used as predictor of outcome remains to be studied.

Another interesting observation is the heterogeneity of the patient responses, with some of them responding very lately more than 25 day post-symptoms. This delayed response might be related to the severity of the infection as some preliminary studies tend to show that pauci-symptomatic patients have lower and delayed antibody response or to the immune status of the patients [8, 12-14]. More systematic clinical and population studies need to be performed to clearly correlate the amplitude and time of the antibody response with i) the severity of the disease, ii) the demographic and clinical data and iii) the immune status of the patients.

The main limitation of this study is that expected positive sera were collected only during the first two month post-symptoms from hospitalized patients presenting moderate to severe symptoms. Our conclusion can thus only be applied to severe clinical presentation and for sera collected during the first two month post-symptoms. Previous studies on other coronavirus [4], or on SARS-CoV-2 [14] suggest that the anti-N antibody response may appeared earlier or simultaneously than the anti-S response and may also waned more rapidly [15]. This implies that the performance of anti-N and anti-S serological tests may be similar during the acute phase of the CoviD-19 disease as this was assessed in this study but that sera from pauci-symptomatic subjects and/or collected more than two month post-exposition may only present a detectable anti-S antibody response.

The anti-N response could be helpful to date the infection since presence of anti-N antibodies would suggest a recent infection. Conversely, anti-S antibody production may be used during all infectious and post-infectious phases, the anti-N and anti-S serological tests being complementary.

This study has identified several SARS-CoV-2 tests exhibiting very good performances of sensitivity and specificity from sera collected from hospitalized patients up to 30-60 days post-symptoms. The SARS-CoV-2 serology may be used as a complement to SARS-CoV-2 RT-PCR for the diagnostic of CoviD-19 and for population sero-prevalence studies. For instance, the serology of SARS-CoV-2 may be used in several clinical and epidemiological assets to confirm or exclude a CoviD-19 disease including i) suggestive clinical symptoms with two consecutive negative RT-PCR, ii) suggestive clinical symptoms with discordant RT-PCRs, iii) infectious control settings for hospitalized patients presenting more than 20 days old suggestive clinical symptoms, iv) CoviD-19 atypical clinical presentations (Guillain-Barré syndrome, meningo-encephalitis, cutaneous vasculitis, Kawasaki disease, diarrhea,…) with negative RT-PCR and v) pre-transplantation or pre-chemotherapy screening [16].

In this study, we have clearly identified robust SARS-CoV-2 anti-N and anti-S serological tests for the diagnosis of patients presenting a moderate to severe CoviD-19 diseases during acute and early sub-acute phase. We demonstrated the limited usefulness of IgM in the serologic diagnostic of SARS-CoV-2. The SARS-CoV-2 serology represents a complementary diagnostic tool to RT-PCR for the CoviD-19 diagnosis on patients presenting both typical and atypical symptoms. Additional evaluation studies of these SARS-CoV-2 anti-N and anti-S serological tests need to be performed to assess the performance of these tests on sera collected more than two month post exposure and from non-hospitalized pauci-symptomatic patients. This is particularly important to assess before conducting large sero-prevalence population studies.

## Data Availability

All data are available on demands

## Author’s contribution

ATC and AC wrote the first draft. All authors critically reviewed the manuscript.

## Ethical statement

This study was evaluated by our Ethics Committee (CER-VD) and they judged that it did not deserve a specific approval being only a quality assessment of diagnostic tests.

## Transparency declaration

The authors have no conflicts of interest to declare.

## Acknowledgement

We warmly thank all the members of the laboratory of serology of the CHUV, Sarah Chappuis, Fabienne Di Paola, Carine Pintér Pont, Claudine Gostely, Emilie Rüegger, Sylvie Caillon-Bouchez, Marijana Vujica, Alexandre Mamin, and Benjamin Gayton, for their wonderful implication in this project, and the set-up of SARS-CoV-2 serologic testing in our hospital. We would like to thank the laboratory of immunology to provide us some sera especially those positive for Lupus, HIV, hepatitis, rheumatoid factors, and auto-antibody. We thank Pr Marchetti Oscar and Pr Pache Antoine from the Ensemble Hospitalier de la Côte, Morges, Switzerland, Dr Caroline Chapuis-Taillard and Dr De Vallière Serge from Clinique de la Source, Lausanne, Switzerland and, Dre Aurélie Jayol-Virely from Clinical laboratory of the Etablissement Hospitalier Nord-Vaudois, Yverdon, Switzerland, to have shared information about the symptoms date of some patients.

